# Cross-reactivity of neutralizing antibody and its correlation with circulating T follicular cells in recovered COVID-19 individuals

**DOI:** 10.1101/2020.06.12.20129460

**Authors:** Jian Zhang, Qian Wu, Ziyan Liu, Qijie Wang, Jiajing Wu, Yabin Hu, Tingting Bai, Ting Xie, Mincheng Huang, Tiantian Wu, Danhong Peng, Weijin Huang, Kun Jin, Ling Niu, Wangyuan Guo, Dixian Luo, Dongzhu Lei, Zhijian Wu, Guicheng Li, Renbin Huang, Yingbiao Lin, Xiangping Xie, Shuangyan He, Yunfan Deng, Jianghua Liu, Weilang Li, Zhongyi Lu, Haifu Chen, Ting Zeng, Qingting Luo, Yi-Ping Li, Youchun Wang, Wenpei Liu, Xiaowang Qu

## Abstract

Seroconversion appeared early after COVID-19 onset, and convalescent sera therapy benefit some critical patients. However, neutralizing antibody (nAb) in convalescents is largely unknown. We found that 97.01% (65/67) of COVID-19 convalescents maintained IgG antibodies with high binding and avidity to SARS-CoV-2 spike subunits S1 and S2, and 95.52% (64/67) had neutralization activity against SARS-CoV-2 pesudovirus, one month after discharge (median ID_50_, 2.75; IQR, 2.34-3.08). Some sera exhibited cross-neutralization against SARS-CoV (76.12%), MERS-CoV (17.91%), or both (10.45%). Interestingly, individuals recovered from severe disease (severe group) had nAbs with binding and neutralization titers higher than non-severe group. Severe group appeared a rapid increase of lymphocytes and a high proportion of circulating CXCR3^+^ Tfh cells. Interestingly, the later were spike-specific and positively correlated with SARS-CoV-2 nAb titers. All subjects had no autoimmunity. Our findings provide novel insights into nAb responses in COVID-19 convalescents and facilitate treatment and vaccine development for SARS-CoV-2 infection.

## Main Text

Coronavirus Disease 2019 (COVID-19), an emerging disease caused by SARS-CoV-2 infection^1-3^, has globally spread causing >4.5 million infections and 0.3 million deaths^4^. Symptoms of COVID-19 range from asymptomatic, mild, moderate to severe^5,6^. No specific drug and vaccine available for COVID-19, and treatments are primarily the supportive cares. Convalescent sera have proven to improve clinical presentation and reduce mortality of critical patients^7,8^. Serum SARS-CoV-2-specific IgM^9-12^ and in most of patients IgG^13-15^ were detectable during 4-to-14 days after symptom onset, though T and B cells decreased dramatically at early acute phase of infection. Few studies reported that neutralizing antibody (nAb) responses to viral spike protein were variable in recovered COVID-19 individuals^1,16,17^. Monoclonal nAbs have been isolated from COVID-19 convalescents and are facilitating clinical trials of antibody therapy and vaccine development^18,19^. However, nAb responses in COVID-19 convalescents are largely unclear, in respect to neutralizing activity, avidity, and cross-neutralization with other coronaviruses. Although lymphocytes play essential role in antibody initiation and maturation, the association of T cell populations with SARS-CoV-2-specific nAb responses remains unknown.

In this study, we recruited 67 recovered COVID-19 individuals. Blood was drawn on day 28 after discharge. The baseline clinical characteristics and laboratory findings on admission was retrospectively analyzed (Extend Data Tables 1 and 2). The binding titer and avidity of serum IgG against S1 and S2 subunits of SARS-CoV-2 spike antigen were examined by ELISA assays (Methods, Online content). All recovered patients had robust anti-S1 (median, 4.61; IQR, 4.01-4.61) and anti-S2 (median, 4.91; IQR, 4.61-5.52) IgG antibodies, and anti-S2 titers were significantly higher than anti-S1 (p<0.001) (Fig. 1a and Extend Data Table 3). All 67 subjects showed avidity index to S1 (median, 44.5; IQR, 34.5-51.75) and S2 (median, 58; IQR, 49-67) antigens (3 samples with marginal binding), and avidity indices to S2 were higher (p<0.001) (Fig.1b), in consistence with the endpoint titers (Fig. 1a). Next, we tested the antibody binding to spike proteins from both SARS-CoV and MERS-CoV. The results showed that 38.81% (26/67) and 73.13% (49/67) had cross-reactions with S1 of SARS-CoV and S2 of MERS-CoV, but not with S1 of MERS-CoV (Fig. 1c). No binding to SARS-CoV S1, and only 6.67%, 1.67% sera from 60 healthy controls showed binding to MERS-CoV S1 and S2, respectively (Extend Data Table 4). We did not test S2 of SARS-CoV due to no qualified antigen available.

**Fig. 1.**
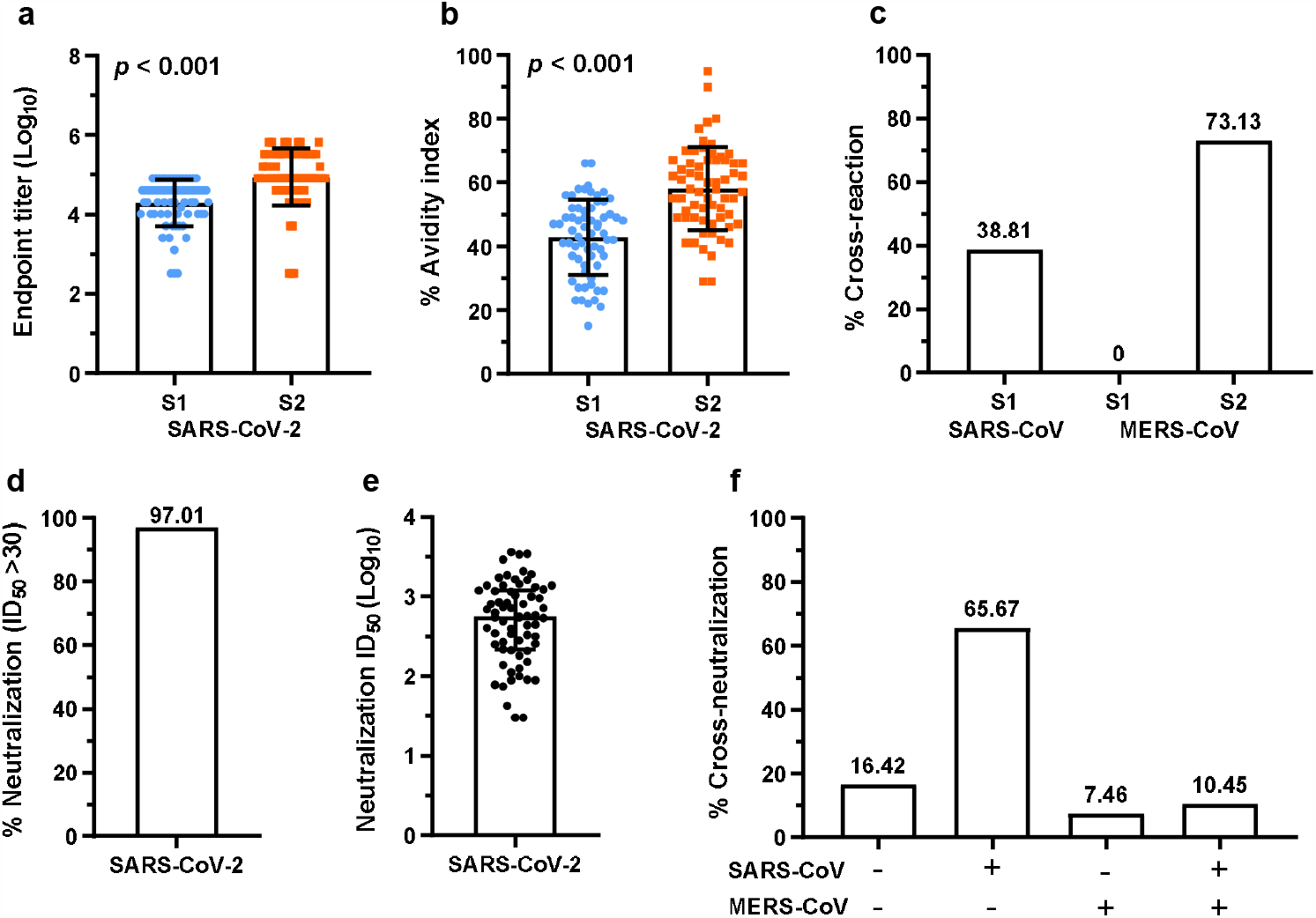
Neutralizing antibody responses and cross-neutralization activity of serum IgG antibodies from recovered COVID-19 individuals. **a**, Endpoint titers of serum IgG antibodies from recovered COVID-19 individuals (n=67) determined by binding to S1 and S2 subunits of SARS-CoV-2 spike protein; Binding to S2 was significantly higher than binding to S1. **b**, Avidity index of serum IgG antibody to S1 and S2 antigens of SARS-CoV-2 (n=64); S2 showed higher avidity. **c**, Percentage of COVID-19 convalescent sera cross-binding with S1 of SARS-CoV spike protein and S1 and S2 of MERS-CoV spike (total, n=67). **d**, Percentage of sera with neutralizing IgG antibody against SARS-CoV-2 pseudovirus particles (64/67, n=67). **e**, Neutralizing titer of sera IgG antibody (n=67) against SARS-CoV-2 pseudovirus particles (ID_50_, Log_10_). **f**, Percentage of serum IgG antibodies (n=67) cross-neutralizing pesudoviruses of SARS-CoV and MERS-CoV; plus “+” and minus “-” stand for positive and negative neutralization, respectively. Endpoint titer and neutralization titer were logarithmic transformed (**a** and **e**). Mann-Whitney U test was used to compare difference between two groups (**a** and **b**), and adjusted *p* value <0.05 was considered significant difference between groups.

To determine the neutralizing and cross-neutralizing activities, we developed SARS-CoV-2 spike-based pesudovirus for neutralization experiments, in parallel with previously developed SARS-CoV and MERS-CoV pesudoviruses^20^. Of 67 individuals, 65 (97.01%) elicited nAbs neutralizing SARS-CoV-2 pesudovirus (median ID_50_, 2.75; IQR 2.34-3.08) (Fig. 1d and e), and neutralization titers were positively correlated with the endpoint titers of SARS-CoV-2 S1- and S2-specific antibodies (Extend Data Fig. 1a-c). Interestingly, 51 (76.12%, 65.67% plus 10.45%) sera showed cross-neutralization with SARS-CoV, 12 (17.91%, 7.46% plus 10.45%) with MERS-CoV, and 7 (10.45%) with both (Fig. 1f). Sera with and without cross-neutralization activity had no differences in neutralization titers for SARS-CoV-S2 (Extend Data Fig. 2a and b), but cross-neutralization activity for SARS-CoV and MERS-CoV were significantly weaker than for SARS-CoV-2 (Extend Data Fig. 2c-e). These findings demonstrate that majority of recovered COVID-19 individuals elicited and maintained robust nAb responses to SARS-CoV-2. Some antibodies had cross-binding and neutralizing activities against SARS-CoV and/or MERS-CoV. No autoimmune antibodies were detected in recovered COVID-19 individuals (Extend Data Table 5).

**Fig. 2.**
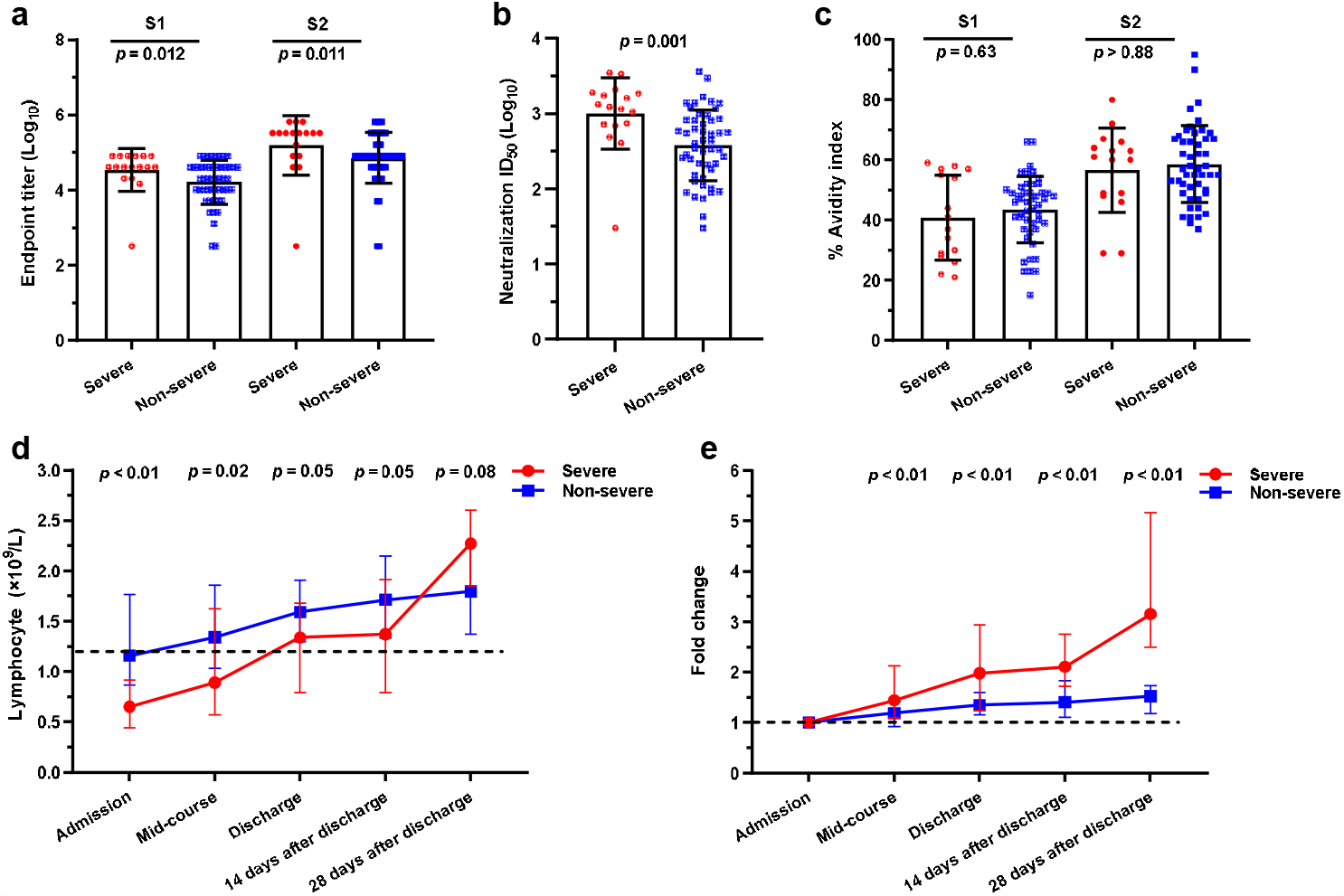
Distinct neutralizing antibody responses and lymphocyte kinetics between subjects recovered from severe and non-severe COVID-19. **a**, Endpoint titers of serum IgG antibodies specific to S1 and S2 of SARS-CoV-2 spike for severe (n=17) and non-severe (n=50) groups; severe group was significantly higher than non-severe group. **b**, Neutralization titers of sera, from severe (n=17) and non-severe (n=50) groups, against SARS-CoV-2 pesudovirues; Titers of severe group was higher than non-severe group. **c**, Avidity of serum IgG antibodies to S1 and S2 of SARS-CoV-2 for severe (n=16) and non-severe groups (n=48); no significant difference between two groups. **d-e**, Kinetics (**d**) and fold change (**e**) of lymphocyte count in severe (red line) and non-severe (blue line) groups; lymphocyte counts of severe group increased faster than non-severe group during the course of disease and after discharge; patient number included in data collection and analysis were the following (severe *vs*. non-severe): on admission (17 *vs*. 50), mid-course of disease (17 *vs*. 41), discharge (16 *vs*. 30), 14 days after discharge (13 *vs*. 40), and 28 days after discharge (13 *vs*. 40). Endpoint titers and neutralization titers were logarithmic transformed (**a** and **b**). Mann-Whitney U test was used to compared difference between two groups (**a-e**), and *p* value <0.05 was considered significant difference between groups.

Patients with severe and mild COVID-19 symptoms were reported with distinct clinical and immunological presentations^9-11^. Here, we categorized all 67 recovered individuals into “severe” (17/67) and “non-severe” (50/67) groups, according to the severity of disease that patients had suffered (Extend Data Table 1). Compared to non-severe group, severe group exhibited higher titers of anti-S1 (p=0.012) and anti-S2 antibodies (p=0.011) (Fig. 2a) and higher neutralization titers (p=0.001) (Fig. 2b), but no difference for binding avidity (Fig. 2c). Next, we analyzed the factors that may associate with nAbs responses and found that nAb response was correlated with the severity of disease (Table 1), though older age (p<0.001), longer course of disease (p=0.007), more comorbidities (p=0.044), and underlying diseases (p<0.001) were higher in severe group (Extend Data Table 1). Further analysis of lymphocyte counts at five time-points, including admission, mid-course, discharge, day 14 and 28 after discharge revealed that majority 16/17 (94.12%) of severe group exhibited lower lymphocyte count on admission, in line with a recent report^5^, while non-severe group that was at the bottom line of normal range (1.2-3.5×10^9^/L) (Fig. 2d). Unlike slow increase in non-severe group, lymphocyte counts of severe group increased gradually during hospitalization and restored to normal on discharge (median, from 0.65 to 2.28×10^9^/L), maintained at this level for two weeks, then unexpectedly rebound from day 14 to 28 after discharge, crossing and having mean value higher than that of non-severe group (p=0.08) (Fig. 2d). Severe group had higher fold change of lymphocytes, related to admission respectively, at all later time points (p<0.01) (Fig. 2e).

**Table 1.**
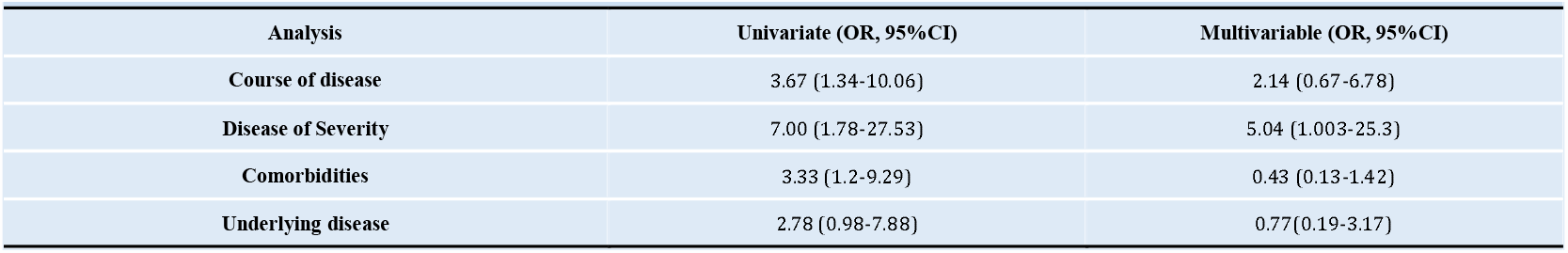
Univariate and multivariable analyses for factor association with neutralizing antibody responses (OR, 95% CI) Data expressed as Odds ratios (OR) and 95% two-sided confidence interval (95% CI). Univariate and Multivariable Binary Logistic Regression models were used to evaluate the influence factors for neutralizing antibody. The median of cutoff dependent variable value (ID_50_, Log_10_) for neutralization were set at 2.41, thus the estimated probability of <2.41 was classified as low neutralization ability group, and a probability of >2.41 was classified as high neutralization ability group. All of covariates were classified by median and changed to two categories of variables for Logistic Regression analysis. Course of disease was defined as the duration (days) from disease onset to discharge.

| Table 1 Analysis of impact factors on neutralization antibody (OR, 95%CI) |  |  |
| --- | --- | --- |
| Analysis | Univariate (OR, 95%CI) | Multivariable (OR, 95%CI) |
| Course of disease | 3.67 (1.34-10.06) | 2.14 (0.67-6.78) |
| Disease of Severity | 7.00 (1.78-27.53) | 5.04 (1.003-25.3) |
| Comorbidities | 3.33 (1.2-9.29) | 0.43 (0.13-1.42) |
| Underlying disease | 2.78 (0.98-7.88) | 0.77(0.19-3.17) |

Next, we investigated whether the increased lymphocytes contributed to nAbs production. Circulating T follicular helper (Tfh) cells, counterparts of germinal center Tfh cells^21^, are important for maturation and antibody production of T-dependent B cells^22,23^. We found that severe and non-severe groups had no difference in circulating Tfh frequency (Fig. 3a), however, CXCR3^+^ Tfh cells (p=0.016) and CXCR3^+^/CXCR3^-^ Tfh cells (p=0.014) were higher in severe group (Fig. 3b-d). Interestingly, nAb titers were positively correlated with CXCR3^+^ Tfh frequency (r=0.486, p=0.012) and the ratio of CXCR3^+^/CXCR3^-^ Tfh cells (r=0.467, p=0.016) (Fig. 3e and f), but not with total Tfh (r=-0.004, p=0.985) and CXCR3^-^ Tfh cells (r=-0.435, p=0.025) (Fig. 3g and h). These findings indicate that CXCR3^+^ Tfh cells may play an important role in supporting or maintaining nAb responses in recovered COVID-19 individuals. Then, we proceeded to examine whether these CXCR3^+^ Tfh cells were SARS-CoV-2 spike-specific. We stimulated PBMCs from convalescents with both S1 and S2 (S1+S2) of SARS-CoV-2 spike and identified spike(S1+S2)-specific proliferation of CD25^+^OX40^+^ CXCR3^+^ Tfh cells (Fig. 3i). We previously found a positive correlation between CXCR3^+^ Tfh cells with HCV nAb titer^24^. Similarly, CXCR3^+^ICOS^+^CXCR5^+^CD4^+^ T cells^25^ and circulating Th1-biased helper cells^26^ were positively associated with influenza virus- and HIV-specific antibody responses, respectively. Together, we concluded that CXCR3^+^ Tfh cell-contribute to initiate and/or maintain nAbs in recovered COVID-19 individuals.

**Fig. 3.**
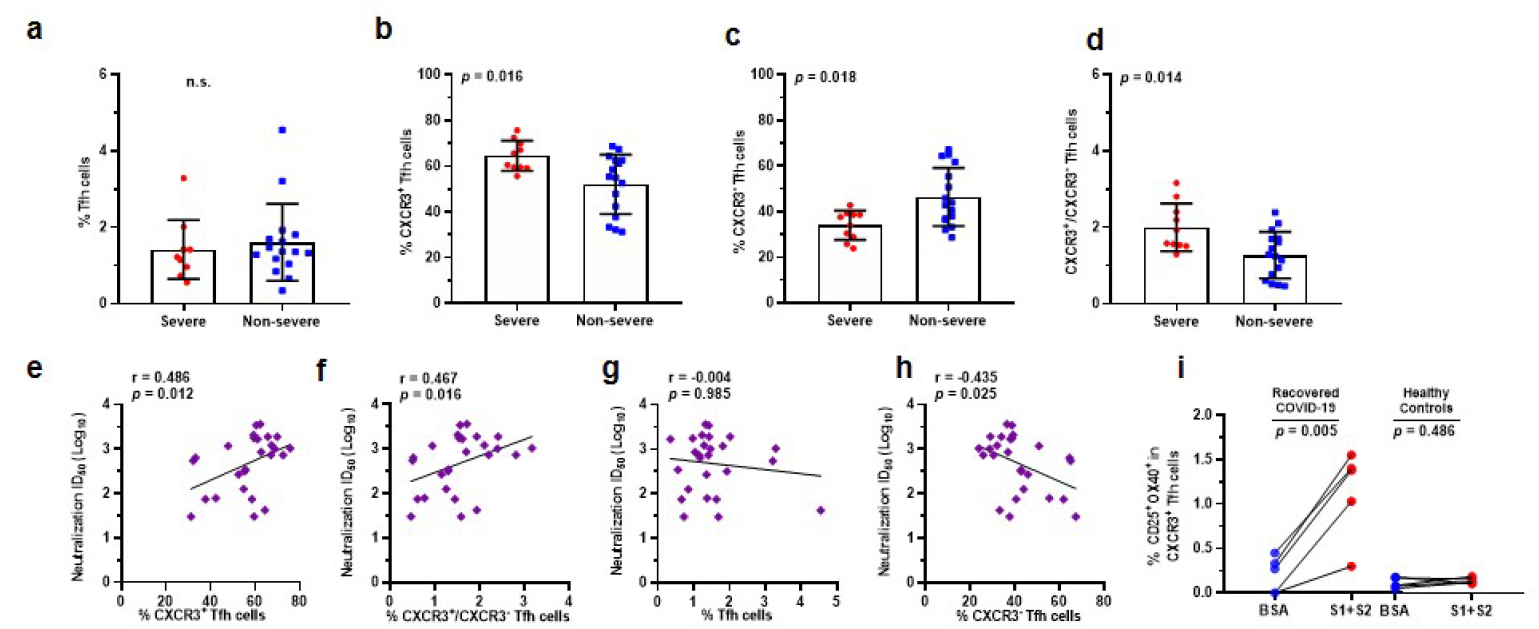
Neutralizing antibody titers were associated with frequencies of circulating CXCR3^+^ Tfh cells in recovered COVID-19 individuals. **a-d**, Frequencies of circulating Tfh cells (Tfh) (**a**), CXCR3^+^ and CXCR3^-^ Tfh subsets (**b, c**), and ratio of CXCR3^+^/CXCR3^-^ Tfh cells (**d**) in severe (n=10) and non-severe (n=16) groups; severe group had a high level of CXCR3^+^ Tfh cells. **e-h**, Correlation analyses of serum neutralization titers and frequencies of CXCR3^+^ Tfh cells (**e**), ratio of CXCR3^+^/CXCR3^-^ Tfh cells (**f**), total Tfh cell cells (**g**), and CXCR3^-^ Tfh subsets (**h**) in recovered COVID-19 individuals (n=26). **i**, Spike-specific circulating Tfh cell response after antigen stimulation. PBMCs from recovered individuals (n=5) and healthy controls (n=5) were stimulated with both S1 and S2 proteins (S1+S2) for 24 hours, antigen-specific Tfh (OX40^+^CD25^+^CXCR3^+^Tfh) cells were analyzed by flow cytometry. Neutralization titers were logarithmic transformed (**e-h**). Mann-Whitney U test was used to compared difference between two groups (**a-d**), Paired t test was used to compared difference between two groups (**i**). Spearman’s rank correlation coefficient was used to describe the relationship between neutralization titers and frequencies of Tfh cells and subsets (**e-h**). *p* value <0.05 was considered significant difference between groups.

We have systematically investigated antibody responses in recovered COVID-19 individuals. Of 67 subjects, 97.01% (65/67) elicited nAbs potently neutralized SARS-CoV-2 (Fig. 1d), of which some had cross-neutralization against SARS-CoV and/or MERS-CoV (Fig. 1f). Variations in cross-neutralization titers between these coronaviruses may be may explained, in part, by sequence homology of spike protein, in which SARS-CoV-2 spike shares ∼76% with SARS-CoV spike but only ∼24% with MERS-CoV spike^27^. Neutralization effects were supported by the endpoint titers and binding avidity against SARS-CoV-2, SARS-CoV, and MERS-CoV (Fig. 1a and b, Extend Data Table 3). nAbs in COVID-19 convalescents was reported but only for those with mild symptoms^1,16^. Cross-binding activity of COVID-19 sera was recently tested against SARS-CoV (spike and S1) and MERS-CoV spike, but only included 3 sera^28^. Patient-derived SARS-CoV monoclonal and RBD-specific CR3022 antibodies could cross-neutralize SARS-CoV-2^29,30^, however low titer of SARS-CoV sera was found without cross-neutralization for SARS-CoV-2^17^. Thus, low levels of nAbs may result in inefficient cross-neutralization. Indeed, antibody titers had positive correlation with neutralization activity (Extend Data Fig. 1a-c). SARS-CoV-2 spike (S1 and S2)-specific nAbs had endpoint titers up to 1:10^4.2-4.4^ dilutions, which may account for the 97.01% neutralization to SARS-CoV-2, and 76.12% (65.67% plus 10.45%) and 17.91% (7.46% plus 10.45%) cross-neutralization to SARS-CoV and MERS-CoV, respectively (Fig. 1f). Cross-neutralization with MERS-CoV was most likely mediated by S2 binding only (Fig. 1c and Extend Data Table 3). Low cross-binding to MERS-CoV spike, but not S1, were recently described using 3 COVID-19 sera^28^. Thus, some COVID-19 patients could elicit nAbs cross-neutralizing MERS-CoV. Interestingly, 10.45% sera cross-neutralized both SARS-CoV and MERS-CoV (Fig. 1f), supporting the cross-neutralization of MERS-CoV by 25% SARS-CoV srea^31^. Existence of cross-neutralizing antibodies offers a possibility to isolate or develop antibodies with neutralizing activity across different coronaviruses.

It is surprising that individuals recovered from severe COVID-19 elicited and maintained higher antibody and neutralization titers than non-severe group (Fig. 2), and the nAb titers positively correlated with severity of disease other than other factors (Table 1, Extend Data Table 1). Similar to what seen for recovered MERS-CoV patients, levels of nAbs positively associated with days in ICU, viral shedding, and ventilation need, several characteristics of critical conditions^32^. We found that SARS-CoV-2 convalescents exhibited low lymphocyte count during the course of disease, but with an accelerated increase, followed by a rebound after discharge (Fig. 2d-e). Rapidly increased lymphocyte counts may be responsible for the high levels of nAb. In recovered MERS-CoV and SARS-CoV-2 patients, nAbs were correlated with antigen-specific CD4^+^ T cells^16,32^, and nAb responses were more stable and longer in recovered severe MERS-CoV patients^33^. Importantly, we found that levels of CXCR3^+^ Tfh cells was significantly higher in recovered severe group than in non-severe group, correlated with nAb responses, and importantly were spike-specific (Fig. 2e-i). Correlations of CXCR3^+^ Tfh cells with nAb responses also exists in other virus infections, such as HCV ^24^ and HIV ^26^. Study on one non-severe COVID-19 patient showed that PD-1^+^ICOS^+^ Tfh cells progressively increased from day 7 after onset of illness^34^. Taken together, circulating CXCR3-biased Tfh cells increased rapidly in severe COVID-19 patients play a critical role in eliciting spike-specific antibodies, with neutralizing activity for SARS-CoV-2 and, to a less extent, for SARS-CoV and MERS-CoV.

In conclusion, we demonstrated that majority of recovered COVID-19 individuals elicited and maintained robust nAb responses, of which some can cross-neutralize SARS-CoV and/or MERS-CoV. The nAb responses are positively correlated with severity of disease and with spike-specific circulating CXCR3^+^ Tfh cells, which were more rapidly populated in recovered severe patients. Beyond this cross-sectional study, the longevity and affinity maturation of nAbs warrant future investigations. Our findings provide important evidence on humoral responses to SARS-CoV-2 infection.

## Data Availability

we stated all the all data referred to this manuscript are available according the below links

## Acknowledgements

We thank all of the participants. This work was supported by the Special Project for Novel Coronavirus of The First People’s Hospital of Chenzhou (X.Q.) and by the SC1-PHE-CORONAVIRUS-2020: Advancing knowledge for the clinical and public health response to the 2019-nCoV epidemic” from European Commission (CORONADX, no. 101003562) (Y.-P.L).

## Author contributions

X.Q., W.L, Y.-P.L. and Y.W. contributed to the study design and data interpretation. Q.W., T.X., M.H., D.P., G.L., X.X., S.H., Y.D., J.L., W.L., Z.L., H.C., T.Z., Q.L., contributed to clinical management, patient recruitment, and data collection. J.Z., Q.W., Z.L., J.W., Y.H., T.B., T.W., W.H., K.J., L.N., W.G., D.L., Y.-P.L., contributed to samples processing, assays development, and experiments conduction. J.Z., Q.W., Z.L., T. W., D.L., and R.H. contributed to statistical analysis and data visualization. X.Q., P.L., Y.-P.L. and J.Z. drafted the manuscript. Y.W., X.Q. and Y.-P.L. contributed to critical revision of the manuscript for important intellectual contents. X.Q. and W.L. provided supervision. All authors meet authorship criteria and approve of publication.

## Methods

### Patients and sample collection

Total 67 recovered COVID-19 patients were enrolled in this study, and diagnosis of COVID-19 was performed according to WHO interim guidance. All of patients came to outpatient showed fever or respiratory symptoms. Chest computed tomography (CT) scans identified abnormal pulmonary nodules, and SARS-CoV-2 infection was further confirmed using real-time PCR by the local health authority. All of patients were hospitalized in Department of Infectious Disease, The Centre Hospital of Shaoyang, Hunan province, China, from January 23 to March 2, 2020. The severity of COVID-19 was graded according to the Chinese Management Guideline for COVID-19 (version 6.0). Of 67 patients, 17 were categorized in severe conditions, and 50 were in mild to moderate symptoms (refer to as non-severe in this study). The medical history and the results of physical, hematological, biochemical, radiological, and microbiological analyses were retrospectively evaluated and analyzed. Peripheral blood of the recovered individuals was collected on day 28 after discharge, corresponding to 44 to 52 days after symptom onset. Peripheral blood mononuclear cells (PBMCs) and serum were isolated and frozen in liquid nitrogen at −80°C ultra-low temperature freezers, respectively.

### Antibody endpoint titer assay

The endpoint titer of antibody was determined by measuring the binding index of serum with S1 and S2 subunits of SARS-CoV-2 spike protein using enzyme linked immunosorbent assay (ELISA). In brief, 96-well plates (Corning, NY, USA) were coated with 20 ng/well S1 or S2 subunit proteins in PBS overnight at 4°C. Plates were washed with 0.05% Tween-20 in PBS (PBS-T) for 5 times and then blocked with blocking buffer (2% FBS and 2% BSA in PBS-T) for 30 minutes. Two-fold serial dilutions, started from 1:20 dilution, were added to the plate in triplicate (100 µl/well) and incubated for 1 hour at room temperature. Spike subunits S1- and S2-specific antibodies were detected by using horseradish peroxidase (HRP)-conjugated anti-human IgG and 3,3’,5,5’-tetramethylbenzidine substrate (Thermo Fisher Scientific, Waltham, MA, USA). Healthy sera were used as negative controls, and monoclonal antibody specific for the receptor binding domain (RBD) of SARS-CoV-2 spike protein (anti-RBD/SARS-CoV-2; made in the lab, unpublished data) was used as positive control. Optical density at 450 nm (OD450) was acquired for each reaction, and the OD450 being 3-fold above the cutoff-OD450 value was considered to be positive. Serum cross-reactivity with S1 of SARS-CoV as well as with both S1 and S2 of MERS-CoV were examined using an optimized serum dilution (1:1000). All of proteins (S1 and S2 of SARS-CoV-2; S1 of SARS-CoV; S1 and S2 of MERS-CoV) were purchased (Sino Biological, China).

### Antibody avidity assay

IgG antibody avidity with S1 and S2 subunits from SARS-CoV-2 were measured by a modified 2-step approach described elsewhere^1,2^. In the first step, serum dilutions were optimized to have an OD450 value between 0.5 and 1.5, so that it could ensure a linear measurement of avidity in next step. The second step was an ELISA assay but included an elution procedure of 1M NaSCN. The avidity index of antibody was calculated as OD NaSCN 1M/OD NaSCN 0M × 100%.

### Antibody neutralization assay

Neutralizing activity of sera was determined by the reduction in luciferase expression, as described previously for the HIV pseudovirus neutralization assay^3^. The 50% inhibitory dilution (ID50) was defined as the serum dilution, at which the relative light units (RLUs) were reduced by 50% compared with the control wells without serum. Background RLUs in the control groups were subtracted from all wells. In brief, pseudovirus were incubated with serial dilutions of sera samples (six dilutions 1:30; 1;90; 1:270; 1:810; 1:2430; 1:7290) in duplicate at 37°C for 1 hour. The control wells were included in duplicate. Naïve Huh7 cells were added to each well and incubated in 5% CO_2_ at 37°C for 24 hours. The luminescence was measured, and the ID_50_ values were calculated with non-linear regression, i.e. log (inhibitor) vs. response (four parameters), using GraphPad Prism 8 (GraphPad Software, Inc., San Diego, CA, USA). Cross-neutralization with SARS-CoV and MERS-CoV were determined using same methods.

### Autoantibody detection

Two milliliters of blood were collected from the participants using EDTA anticoagulation tubes (BD Biosciences, Franklin Lake, NJ, USA), and sera were immediately isolated by centrifugation (3,000 rpm) for 5 minutes (Sorvall ST 40R Centrifuge, Thermo Fisher Scientific). Sixteen autoimmune antibodies were tested to examine whether the autoimmunity occurred after recovery from SARS-CoV-2 infection. Of which, anti-dsDNA and anti-ANA antibodies were detected by ELISA (Zeus Scientific, Inc. New Jersey, USA), while anti-nucleosomes, histones, SmD1, U1-SnRNP, SS-A/Ro60KD, SS-A/Ro52 KD, SS-B/La, Sc1-70, CENP-B, Jo-1, and anti-PO/38KD were examined by Line Immuno Assay (LIA), according to the manufacturer’s protocols (HUMAN GmbH, Magdeburg, Germany).

### Flow cytometry

Ten milliliters of blood were collected from recovered COVID-19 patients and heathy volunteers with EDTA anticoagulant tubes (BD Biosciences). PBMCs were immediately isolated by Ficoll density gradient centrifugation, according to the manufacturer’s protocol (GE Healthcare Bio-Sciences AB, Kontaktuppgifter, Sweden), and stored in liquid nitrogen using a programmed cooling procedure.

To analyze the circulating follicular helper T cells (Tfh) and Tfh subsets, cryopreserved PBMCs were thawed at 37°C water bath and cultured immediately in RPMI 1640 medium supplemented with 10% FBS overnight in 5% CO2 at 37°C. For cell surface staining, 1×10^6^ PBMCs/mL were first labeled with a LIVE/DEAD® Fixable Blue Dead Cell Stain Kit (Thermo Fisher Scientific) to exclude dead cells, and then treated with Fc Block (BioLegend, San Diego, CA, USA) to block non-specific binding. The treated PBMCs were stained with antibodies, which had been pre-titrated and fluorescently labeled, in 96-well V-bottom plates at 4°C for 30 minutes. The fluorescently labeled antibodies used in this study were BUV737 mouse anti-human CD4 (SK3) and PE mouse anti-human CXCR3 (1C6) (BD Biosciences), FITC mouse anti-human PD-1 (EH12.2H7) (BioLegend), and PE-eFluor 610 mouse anti-human CXCR5 (MU5UBEE) (Thermo Fisher Scientific, Waltham, MA, USA). Cell population gating was set based on the mean fluorescence intensity “minus one” (FMO) and unstained controls. Samples were loaded onto a MoFlo XDP Flow Cytometer (Beckman Coulter, Brea, CA, USA) immediately after antibody staining. All data were analyzed with FlowJo 10.0 software (Tree Star, San Carlos, CA, USA).

### Antigen-specific Tfh cell assay

Cryopreserved PBMCs were thawed and cultured in complete RPMI1640 medium in 5% CO2 at 37°C incubator overnight. Cells were stimulated with SARS-CoV-2 spike protein (S1 plus S2) (5μg/mL) or negative control BSA (5μg/mL) (Sigma-Aldrich, St. Louis, MO, USA) in 5% CO2 at 37°C for 24 hours. PBMCs from healthy controls were included in parallel as controls. Circulating Tfh (cTfh) cells were gated as live CD4^+^ PD-1^+^ CXCR5^+^ T cells. PE-Cy™5 mouse anti-human CD25 (M-A251) (BD Biosciences) and APC mouse anti-human OX40 (ACT35) (BioLegend) were used to define antigen-specific (CD25^+^ OX40^+^) Tfh cell responses after stimulation as described previously^4,5^.

### Statistical analysis

Baseline clinical characteristics data were non normal distribution, so continuous variables were expressed as median and Interquartile Range (IQR). Rank variables was expressed as constituent ratio. Mann-Whitney U test and χ^2^ or Fisher’s Exact tests were used in the comparison of two different groups. Paired Sample t test was used to compare the antigen-specific Tfh cells before and after the stimulation with S1 and S2 proteins. Kruskal-Wallis test was used for the comparison of multiple groups, and Dunn’s multiple comparisons test was used between two groups. Spearman’s Rank Correlation Coefficient was used to measure the correlation between two variables of Tfh cells, neutralization antibodies, and binding avidity. Univariate and Multivariable Binary Logistic Regression model was used to rank the factors potential associations with nAb responses. Odds ratios (ORs) and 95% two-sided confidence interval (95% CI) were generalized by equation models to describe the factors contributing to nAb responses.

Analyses of the data were done by SPSS version 26 and GraphPad Prism version 8.0.

### Ethics approval

This study was performed in accordance with the Good Clinical Practice and the Declaration of Helsinki principles for ethical research. The study protocol was approved by the Institutional Review Board of The Center Hospital of Shaoyang (V1. 0, 20200301), Hunan Province, China. Written informed consent was obtained from each participant. Medical data were collected from electronic records of the hospitals using standardized Data Collection Forms recommended by the International Severe Acute Respiratory and Emerging Infection Consortium.

## Extend Data Figures and Tables

**Extend Data Figure 1.**
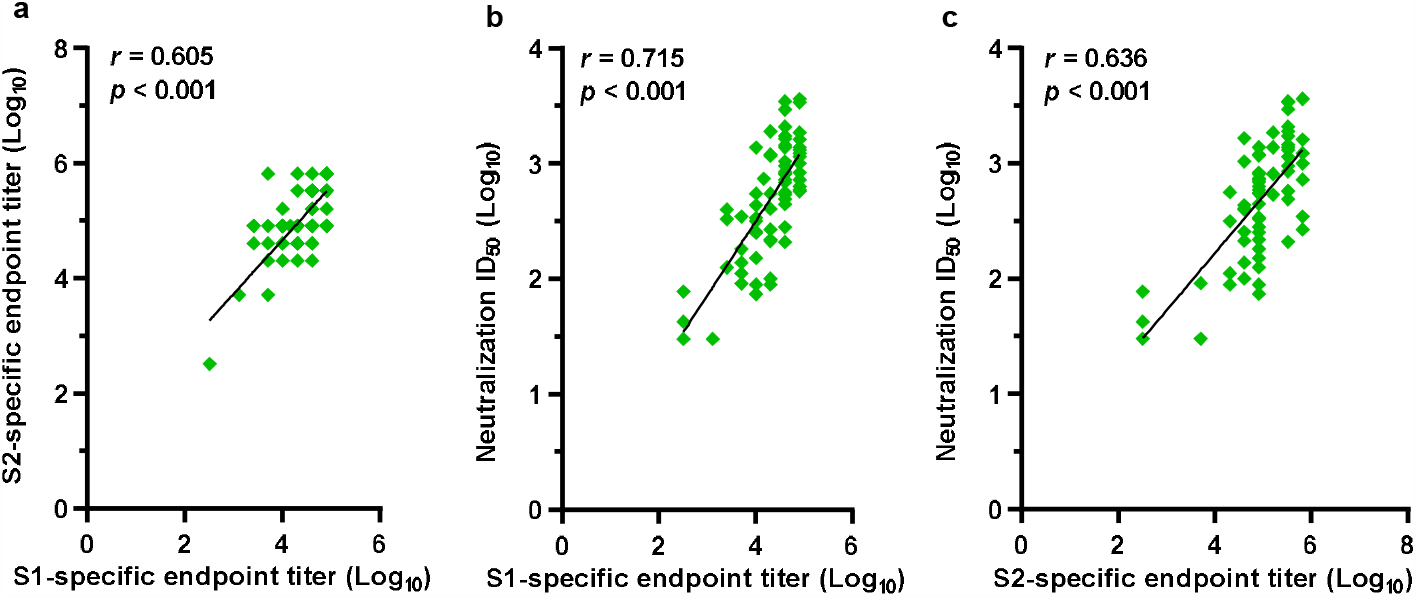
Correlation analysis of the serum neutralization titers and the endpoint titers of anti-S1 and anti-S2 antibodies. Correlation analysis was performed, using Spearman’s rank coefficient of correlation, to identify the strength of relationships between neutralization titers of serum antibodies against SARS-CoV-2 pesudovirus and the endpoint titers (binding activity) of antibodies binding to S1 and S2 subunits of SARS-CoV-2. Correlation between S1- and S2-specific endpoint titers of serum antibodies (**a**), sera neutralization titers and S1-specific endpoint titers (**b**), and sera neutralization titers and S2-specific endpoint titers (**c**); Positive correlations existed between S1- and S2-specific endpoint titers as well as between neutralization and S1- or S2-specific endpoint titers. *p* value <0.05 was considered to be significantly different between groups.

**Extend Data Figure 2.**
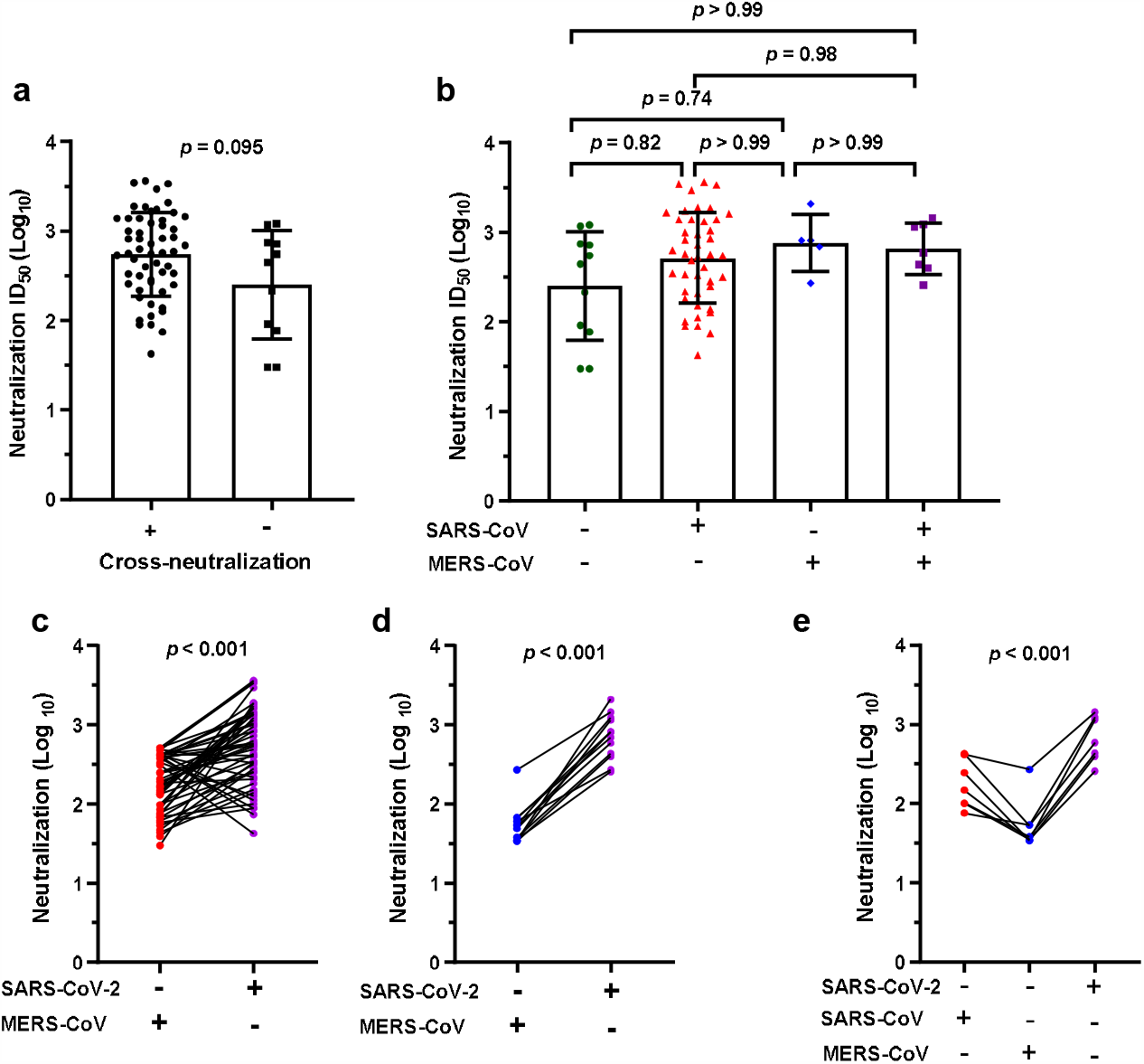
Neutralization titers of SARS-CoV-2 pesudovirus for the serum IgG antibodies that had cross-neutralization with SARS-CoV and MERS-CoV. **a**. Serum neutralizing titers against SARS-CoV-2 pesudovirus were divided into two groups, with and without cross-neutralization activity against SARS-CoV and/or MERS-CoV; No difference between two groups. **b**. Comparison of SARS-CoV-2-specific neutralizing titers of the sera with cross-neutralization with SARS-CoV and/or MERS-CoV; No difference between two groups. **c-e**. Comparison of neutralization titers for SARS-CoV-2 and cross-neutralization titers for SARS-CoV, or for MERS-CoV, for individual COVID-19 convalescent serum; cross-neutralization titers for SARS-CoV or MERS-CoV were lower than that for SARS-CoV-2.

**Extend Data Figure 3.**
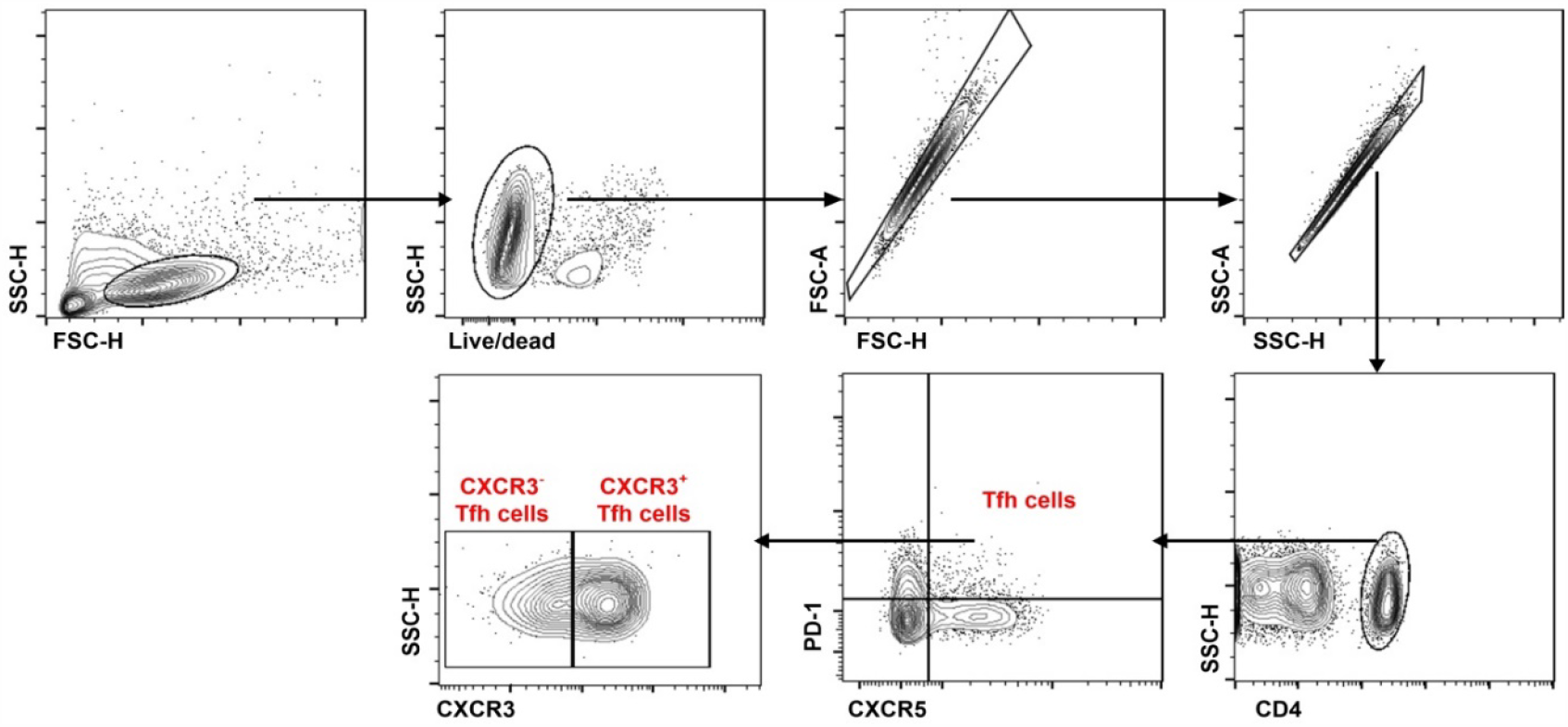
Gating strategy of circulating Tfh cells in recovered COVID-19 individuals. Representative gating strategy for circulating Tfh cells, CXCR3^+^ and CXCR3^-^ Tfh cells in the flow cytometry analysis. Gating was based on the mean fluorescence intensity “minus one” (FMO).

**Extend Data Figure 4.**
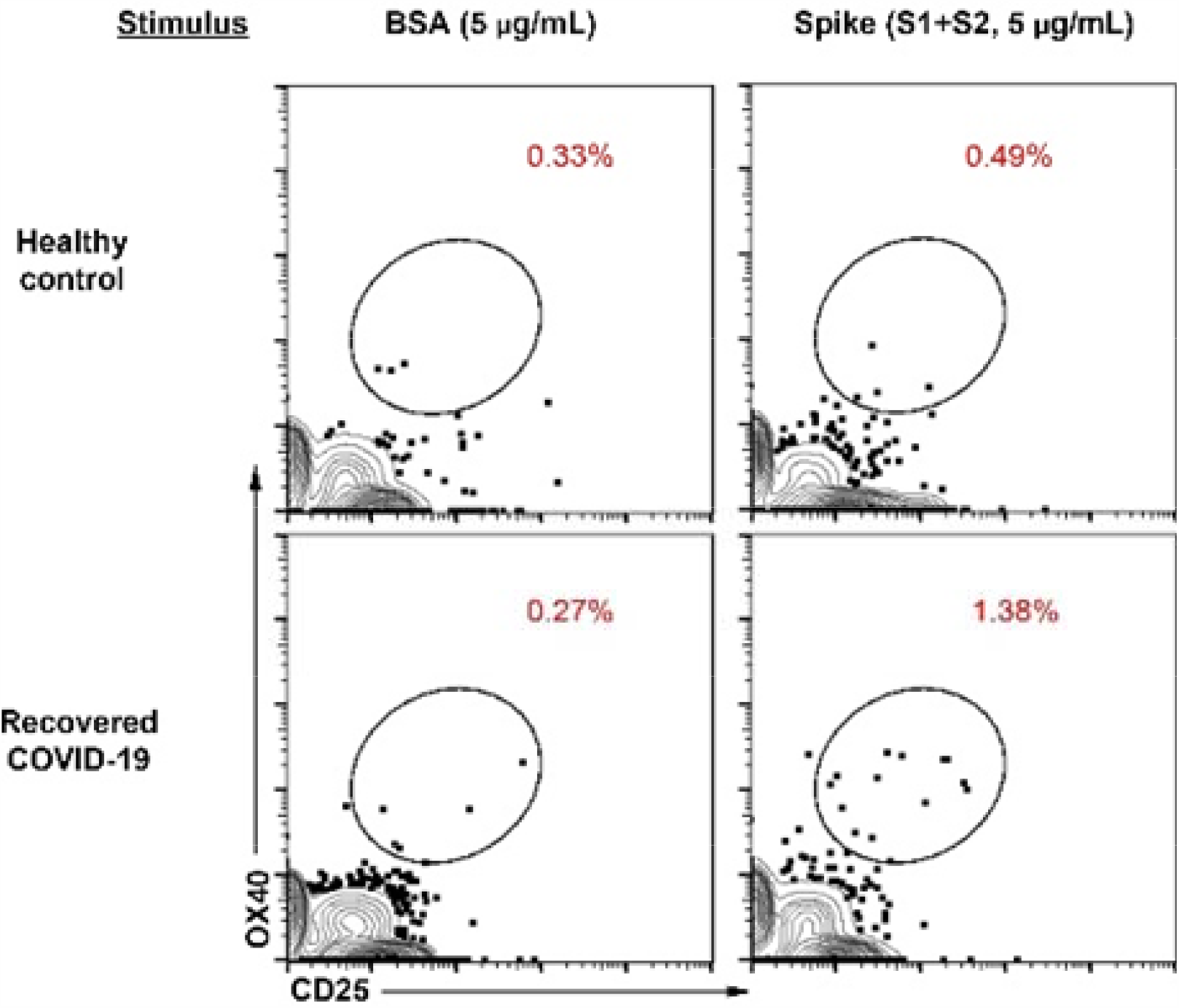
Spike-specific Tfh cell responses after antigen stimulation in vitro. Representative flow cytometry plots for the spike antigen-specific CXCR3^+^ Tfh cell (CD25^+^ OX40^+^ CXCR3^+^ Tfh cells) responses from healthy control and recovered COVID-19 individuals. Gating was based on the mean fluorescence intensity “minus one” (FMO). PBMCs were stimulated with S1 plus S2 subunits (5μg/mL) of SARS-CoV-2 spike protein in 5% CO2 at 37°C for 24 hours. Cells stimulated with BSA (5μg/mL) were negative control.

## Extend Tables

**Extend Data Table 1.** Baseline characteristics of COVID-19 patients recruited in this study.

|  |  |  |  |  |
| --- | --- | --- | --- | --- |
|  | No (%) |  |  |  |
|  | Total (n = 67) | Severe (n=17) | Non-severe (n=50) | P value |
| **Basic Information** |  |  |  |  |
| Age (years) (IQR) | 43(30-53) | 59 (45-69.5) | 41(29-47) | <0.001* |
| Sex |  |  |  |  |
| Male | 34(50.8) | 11(64.7) | 23(46) | 0.26 |
| Female | 33(49.3) | 6(35.3) | 27(54) |  |
| Course of disease (days) (IQR) | 20(16-25) | 25(21-28) | 18(16-23.3) | 0.007* |
| **Presenting symptoms and signs** |  |  |  |  |
| Cough | 59(88.1) | 17(100) | 42(84) | 0.10 |
| Fever | 58(86.6) | 17(100) | 41(82) | 0.10 |
| Fatigue | 41(61.2) | 12(70.6) | 29(58) | 0.40 |
| Expectoration | 28(41.8) | 12(70.6) | 16(32) | 0.009* |
| Dyspnea | 26(38.8) | 15(88.2) | 11(22) | <0.001* |
| Myalgia | 26(38.8) | 6(35.3) | 20(40) | 0.78 |
| Chills | 21(31.3) | 12(70.6) | 9(18) | <0.001* |
| Headache | 16(23.9) | 5(29.4) | 11(22) | 0.53 |
| Anhelation | 10(14.9) | 5(29.4) | 5(10) | 0.11 |
| Diarrhea | 5(7.5) | 2(11.8) | 3(6) | 0.60 |
| Maximum body temperature (°C) (IQR) | 38.3(37.7-38.7) | 38.5(38-39.1) | 38.3(37.5-38.5) | 0.054 |
| **Underlying disease** |  |  |  |  |
| Cardiovascular diseases | 16(23.9) | 9(52.9) | 7(14) | 0.002* |
| Hypertension | 13(19.4) | 8(47.1) | 5(10) | 0.002* |
| Diabetes | 9(13.4) | 6(35.3) | 3(6) | 0.006* |
| COPD | 4(6) | 3(17.7) | 1(2) | 0.047* |
| HBV | 3(4.5) | 1(5.9) | 2(4) | >0.99 |
| Total underlying disease | 24(35.8) | 13(76.5) | 11(22) | <0.001 |
| **Comorbidities** |  |  |  |  |
| Liver insufficiency | 29(43.3) | 13(76.5) | 16(32) | 0.002* |
| Cardiac insufficiency | 25(37.3) | 9(52.9) | 16(32) | 0.15 |
| Renal insufficiency | 4(6) | 2(11.8) | 2(4) | 0.27 |
| Total Comorbidities | 29(43.28) | 14(82.4) | 26(52) | 0.044 |
| Data of continuous variable was expressed as median (IQR), rank variables was expressed as positive cases (percentage). Inequality hypothesis tests were done with $\chi^2$ or Fisher's exact tests for proportions and with Mann-Whitney U test. *$p < 0.05$ was considered to be significant difference between groups. | | | | |

**Extend Data Table 2.** Laboratory findings of patients with COVID-19 on admission.

|  | Median (IQR) |  |  |  |  |
| --- | --- | --- | --- | --- | --- |
|  | Normal range | Total (n = 67) | Severe (n=17) | Non-severe (n=50) | P value |
| Blood routine |  |  |  |  |  |
| White blood cell count (×10<sup>9</sup> l<sup>-1</sup>) | 4-10 | 5.4(4.2-8.7) | 5.4(4.1-11.3) | 5.4(4.1-8.1) | 0.41 |
| Red blood cell count (×10<sup>9</sup> l<sup>-1</sup>) | 3.5-5 | 4.7(4.2-5.1) | 4.7(4.1-5.2) | 4.8(4.2-5.1) | 0.49 |
| Neutrophil count (×10<sup>9</sup> l<sup>-1</sup>) | 1.4-7 | 3.7(2.5-6.9) | 4.7(2.7-8.7) | 3.7(2.3-5.9) | 0.08 |
| Neutrophil percentage (%) | 45-75 | 66(56.9-77) | 63.9(60.2-76.4) | 66.5(55.9-77.4) | 0.83 |
| Lymphocyte count (×10<sup>9</sup> l<sup>-1</sup>) | 1.2-3.5 | 1(0.7-1.7) | 0.7(0.4-0.9) | 1.2(0.9-1.8) | <0.001* |
| Lymphocyte percentage (%) | 20-40 | 23(14.1-31.7) | 24.9(13.5-28.3) | 22.6(14.5-33.9) | 0.67 |
| Monocyte count (×10<sup>9</sup> l<sup>-1</sup>) | 0.1-1.2 | 0.53(0.38-0.63) | 0.49(0.29-0.66) | 0.54(0.39-0.62) | 0.92 |
| Monocyte percentage (%) | 3-12 | 8.5(6.5-11.1) | 8.9(6.9-12.3) | 8.4(6.5-10.9) | 0.35 |
| Platelet count (×10<sup>9</sup> l<sup>-1</sup>) | 125-350 | 223(154-258) | 154(133-191) | 234.5(187-270.8) | 0.001* |
| Hemoglobin (g l<sup>-1</sup>) | 120-160 | 142(126-153) | 137(122-150.5) | 142.5(126-157) | 0.45 |
| Hematocrit (%) | 35.0-51 | 40.1(36.2-44.9) | 38.1(36.1-43.5) | 41(36.1-45.4) | 0.24 |
| Basophil percentage (%) | 0-1 | 0.11(0-0.5) | 0.12(0-0.5) | 0.11(0-0.5) | 0.93 |
| Basophil count (×10<sup>9</sup> l<sup>-1</sup>) | 0.12-08 | 0.01(0-0.04) | 0.01(0-0.03) | 0.01(0-0.04) | 0.64 |
| Eosinophil percentage (%) | 1-5 | 0.5(0-3.2) | 0.4(0-1.7) | 0.5(0-3.2) | 0.78 |
| Eosinophil count (×10<sup>9</sup> l<sup>-1</sup>) | 0.05-0.3 | 0.03(0-0.17) | 0.02(0-0.08) | 0.03(0-0.17) | 0.66 |
| Blood biochemistry |  |  |  |  |  |
| Albumin (g l<sup>-1</sup>) | 40-55 | 40.4(36.1-43.5) | 36.1(33.5-38.6) | 41.7(38.3-45.2) | <0.001* |
| Total protein (g l<sup>-1</sup>) | 62-85 | 68(64-72.6) | 67.8(63-73.7) | 68.3(64.2-72.6) | 0.86 |
| Creatinine (μmol l<sup>-1</sup>) | 40-88 | 62.7(52.3-80.2) | 80.2(63.6-86.4) | 61.2(46.7-77.1) | 0.007* |
| Creatine kinase (U l<sup>-1</sup>) | 25-190 | 70.5(50.8-112.8) | 102(49-169.3) | 68(51.2-96.6) | 0.08 |
| Creatine kinase-MB (U l<sup>-1</sup>) | 0-24 | 15(12.4-20) | 15.5(13-23) | 15(11.9-19.3) | 0.45 |
| Lactate dehydrogenase (U l<sup>-1</sup>) | 109-245 | 238.5(192.1-316.3) | 316(266.5-412.2) | 225.1(184.8-289.8) | <0.001* |
| Alanine aminotransferase (U l<sup>-1</sup>) | 8-45 | 21(14-37) | 28.8(15.4-48.1) | 18.9(13.6-35.3) | 0.31 |
| Aspartate aminotransferase (U l<sup>-1</sup>) | 14-38 | 27(21.3-35) | 31.5(25.2-50.3) | 25.6(20.2-32.1) | 0.019* |
| Total bilirubin (mmol l<sup>-1</sup>) | 2-20.4 | 12.3(8.1-20.3) | 15.9(10-22.2) | 12(7.2-19.9) | 0.31 |
| Total bile acid (μmol l<sup>-1</sup>) | 0-10 | 4.5(1.9-7.8) | 4.5(2.2-8.1) | 4.1(1.7-7.6) | 0.42 |
| Total cholesterol (mmol l<sup>-1</sup>) | 0-5.2 | 3.7(3.1-4.4) | 3.3(2.9-4) | 3.8(3.2-4.4) | 0.08 |
| Urea (mmol l<sup>-1</sup>) | 2-8.3 | 4(3.3-4.9) | 4.6(3.7-5.8) | 3.9(3.1-4.6) | 0.07 |
| Uric acid, (μmol l<sup>-1</sup>) | 150-450 | 249.4(177.2-33) | 208.2(155.3-284.5) | 254.8(186.2-332.9) | 0.09 |
| Potassium, (mmol l<sup>-1</sup>) | 3.5-5.3 | 3.8(3.5-4.1) | 3.6(3.4-4) | 3.9(3.6-4.2) | 0.09 |
| Sodium (mmol l<sup>-1</sup>) | 137-147 | 141.4(138.4-144.5) | 137.1(134.8-142.7) | 142(139.2-145) | 0.016* |
| Glucose (mmol l<sup>-1</sup>) | 3.9-6.1 | 7.1(5.7-9.9) | 9.4(7.7-14.3) | 6.3(5.5-8.5) | 0.001* |
| Myoglobin (μg l<sup>-1</sup>) | 0-90 | 57.4(48.3-71.2) | 58.8(51.7-72.2) | 57.3(47.2-70.9) | 0.81 |
| Alkaline phosphatase (U l<sup>-1</sup>) | 35-125 | 49.4(37.7-76.1) | 86.3(56.4-172.1) | 43.2(35.1-61.3) | <0.001* |
| β2microglobulin (mg l<sup>-1</sup>) | 1-3 | 2.7(2.1-3.8) | 3.2(2.5-4.1) | 2.6(2-3.6) | 0.06 |
| Procalcitonin (ng ml<sup>-1</sup>) | 0-0.05 | 0.03(0-0.05) | 0.04(0.02-0.09) | 0.03(0-0.05) | 0.031* |
| Erythrocyte sedimentation rate (mm h<sup>-1</sup>) | 0-15 | 42(18.5-80.5) | 73(22.6-93.7) | 39(16-56) | 0.12 |
| Coagulation function |  |  |  |  |  |
| Prothrombin time (s) | 8-14 | 11.1(10.4-12) | 11.3(10.5-12) | 11(10.3-12.1) | 0.67 |
| Thrombin time (s) | 14-21 | 17.3(16.4-17.8) | 16.6(16.2-17.7) | 17.3(16.5-18) | 0.16 |
| Activated partial thromboplastin time (s) | 20-40 | 30.2(26.3-37) | 33.5(30.1-39.7) | 29(23.1-36.7) | 0.021* |
| D-dimer (mg l<sup>-1</sup>) | 0-0.55 | 0.29(0.16-0.47) | 0.48(0.28-0.7) | 0.21(0.16-0.43) | 0.003* |

All of these data were collected on admission. Data were expressed as median (IQR), and inequality hypothesis tests were done with Mann-Whitney U test. \* $p < 0.05$ was considered to be significant difference between groups.

**Extend Data Table 3.** Anti-SARS-CoV-2 spike antibody responses in recovered COVID-19 patients.

| Patient ID | Endpoint titer |  | Avidity index (%) |  | Cross-reaction |  |  | Neutralization (ID<sub>50</sub>) |  |  |
| --- | --- | --- | --- | --- | --- | --- | --- | --- | --- | --- |
|  | SARS-CoV-2 |  | SARS-Cov-2 |  | SARS-CoV | MERS- CoV |  | SARS-CoV-2 | SARS-CoV | MERS-CoV |
|  | S1 | S2 | S1 | S2 | S1 | S1 | S2 |  |  |  |
| Severe (n=17) |  |  |  |  |  |  |  |  |  |  |
| PT1 | 14960 | 81920 | 44 | 48 | + | - | + | 735 | <30 | <30 |
| PT2 | 40960 | 655360 | 28 | 80 | + | - | + | 728 | 253 | <30 |
| PT3 | 40960 | 327680 | 22 | 46 | + | - | + | 2067 | <30 | 35 |
| PT4 | 40960 | 40960 | 54 | 63 | + | - | + | 1037 | 406 | <30 |
| PT5 | 40960 | 81920 | 59 | 63 | + | - | + | 686 | <30 | 68 |
| PT6 | 81920 | 327680 | 57 | 67 | + | - | + | 3397 | 476 | <30 |
| PT7 | 40960 | 327680 | 29 | 29 | + | - | + | 1750 | 73 | <30 |
| PT8 | 81920 | 327680 | 57 | 60 | + | - | - | 1316 | 518 | <30 |
| PT9 | 81920 | 655360 | 58 | 61 | + | - | - | 1625 | 98 | <30 |
| PT10 | 81920 | 163840 | 41 | 72 | - | - | + | 1881 | 328 | <30 |
| PT11 | 320 | 320 | NA | NA | - | - | - | <30 | <30 | <30 |
| PT12 | 40960 | 327680 | 21 | 66 | - | - | + | 3457 | 513 | <30 |
| PT13 | 81920 | 655360 | 26 | 60 | - | - | + | 1225 | 76 | 54 |
| PT14 | 20480 | 327680 | 34 | 49 | - | - | + | 1898 | 30 | <30 |
| PT15 | 20480 | 40960 | 55 | 49 | - | - | + | 403 | 426 | <30 |
| PT16 | 40960 | 327680 | 30 | 29 | - | - | + | 495 | 249 | <30 |
| PT17 | 81920 | 327680 | 37 | 64 | - | - | + | 1149 | 149 | 35 |
| Non-severe (n=50) |  |  |  |  |  |  |  |  |  |  |
| PT18 | 20480 | 81920 | 37 | 70 | + | - | + | 218 | 409 | <30 |
| PT19 | 81920 | 81920 | 47 | 66 | + | - | + | 631 | 209 | <30 |
| PT20 | 40960 | 327680 | 44 | 77 | + | - | + | 859 | 160 | <30 |
| PT21 | 81920 | 81920 | 47 | 62 | + | - | + | 722 | <30 | <30 |
| PT22 | 81920 | 81920 | 52 | 58 | + | - | + | 585 | 423 | 53 |
| PT23 | 81920 | 655360 | 40 | 41 | + | - | + | 1006 | 398 | <30 |
| PT24 | 40960 | 40960 | 49 | 53 | + | - | + | 1673 | 133 | <30 |
| PT25 | 40960 | 20480 | 27 | 39 | + | - | - | 558 | 191 | <30 |
| PT26 | 81920 | 655360 | 37 | 49 | + | - | - | 3615 | 483 | <30 |
| PT27 | 20480 | 81920 | 40 | 55 | + | - | + | 1168 | <30 | <30 |
| PT28 | 20480 | 81920 | 45 | 41 | + | - | + | 1214 | <30 | <30 |
| PT29 | 20480 | 655360 | 26 | 70 | + | - | + | 269 | <30 | 61 |
| PT30 | 40960 | 163840 | 48 | 62 | + | - | + | 815 | <30 | 49 |
| PT31 | 81920 | 327680 | 47 | 55 | + | - | + | 576 | 376 | <30 |
| PT32 | 81920 | 327680 | 49 | 47 | + | - | + | 1395 | 141 | <30 |
| PT33 | 40960 | 327680 | 23 | 55 | + | - | + | 956 | 159 | <30 |
| PT34 | 40960 | 327680 | 15 | 58 | + | - | + | 2941 | 86 | <30 |
| PT35 | 20480 | 40960 | 52 | 68 | - | - | + | 100 | 70 | <30 |
| PT36 | 10240 | 81920 | 39 | 55 | - | - | - | 75 | 394 | <30 |
| PT37 | 2560 | 81920 | 66 | 90 | - | - | + | 332 | 486 | <30 |
| PT38 | 1280 | 5120 | 42 | 69 | - | - | - | <30 | <30 | <30 |
| PT39 | 5120 | 655360 | 58 | 95 | - | - | + | 344 | 136 | <30 |
| PT40 | 10240 | 20480 | 52 | 69 | - | - | + | 316 | 68 | <30 |
| PT41 | 20480 | 20480 | 32 | 49 | - | - | - | 90 | 380 | <30 |
| PT42 | 10240 | 81920 | 46 | 67 | - | - | + | 544 | <30 | <30 |
| PT43 | 320 | 320 | NA | NA | - | - | - | 79 | <30 | <30 |
| PT44 | 10240 | 40960 | 41 | 41 | - | - | + | 255 | 99 | 36 |
| PT45 | 20480 | 81920 | 49 | 73 | - | - | + | 546 | 390 | <30 |
| PT46 | 40960 | 81920 | 48 | 55 | - | - | + | 282 | 39 | <30 |
| PT47 | 2560 | 81920 | 54 | 79 | - | - | + | 125 | 337 | <30 |
| PT48 | 320 | 320 | NA | NA | - | - | + | 42 | 309 | <30 |
| PT49 | 20480 | 40960 | 34 | 57 | - | - | + | 215 | <30 | <30 |
| PT50 | 40960 | 81920 | 23 | 52 | - | - | - | 1395 | 169 | <30 |
| PT51 | 40960 | 327680 | 39 | 57 | - | - | + | 211 | 185 | <30 |
| PT52 | 40960 | 81920 | 41 | 52 | - | - | + | 813 | <30 | 67 |
| PT53 | 10240 | 81920 | 49 | 67 | - | - | - | 249 | 77 | <30 |
| PT54 | 40960 | 81920 | 51 | 42 | - | - | - | 452 | <30 | <30 |
| PT55 | 40960 | 163840 | 27 | 44 | - | - | + | 540 | 188 | <30 |
| PT56 | 40960 | 327680 | 23 | 37 | - | - | + | 1433 | 420 | 272 |
| PT57 | 2560 | 40960 | 49 | 51 | - | - | + | 395 | 248 | 34 |
| PT58 | 5120 | 20480 | 56 | 61 | - | - | - | 113 | 440 | <30 |
| PT59 | 10240 | 163840 | 56 | 53 | - | - | + | 1389 | 416 | <30 |
| PT60 | 5120 | 5120 | 66 | 65 | - | - | - | 92 | <30 | <30 |
| PT61 | 81920 | 81920 | 43 | 67 | - | - | - | 837 | 420 | <30 |
| PT62 | 10240 | 81920 | 56 | 62 | - | - | + | 335 | 47 | <30 |
| PT63 | 5120 | 81920 | 42 | 44 | - | - | + | 184 | 62 | <30 |
| PT64 | 5120 | 40960 | 48 | 71 | - | - | - | 139 | 44 | <30 |
| PT65 | 10240 | 81920 | 36 | 47 | - | - | + | 88 | 59 | <30 |
| PT66 | 10240 | 81920 | 50 | 66 | - | - | - | 151 | 54 | <30 |
| PT67 | 10240 | 40960 | 46 | 50 | - | - | - | 436 | 101 | 38 |

Antibody binding titers to S1 and S2 of SARS-CoV-2 spike were performed by endpoint dilution assay and expressed as endpoint dilution. Cross-reactivity with SARS-CoV S1 and MERS-CoV S1 and S2 proteins were expressed positive (+) or negative (-). Neutralization (SARS-CoV-2) and cross-neutralization (SARS-CoV and MERS-CoV) were expressed as ID<sub>50</sub>, ID<sub>50</sub> >30 was defined as positive according to the cutoff value of neutralization assay.

**Extend Data Table 4.** SARS-CoV, MERS-CoV spike-specific antibody responses in healthy controls.

| Healthy control | SARS-CoV S1 | MERS-CoV S1 | MERS-CoV S2 |
| --- | --- | --- | --- |
| HC1 | - | - | - |
| HC2 | - | - | - |
| HC3 | - | - | - |
| HC4 | - | - | - |
| HC5 | - | - | - |
| HC6 | - | + | - |
| HC7 | - | - | - |
| HC8 | - | - | - |
| HC9 | - | - | - |
| HC10 | - | - | - |
| HC11 | - | - | - |
| HC12 | - | - | - |
| HC13 | - | - | - |
| HC14 | - | - | - |
| HC15 | - | - | - |
| HC16 | - | - | - |
| HC17 | - | - | - |
| HC18 | - | - | - |
| HC19 | - | - | - |
| HC20 | - | - | - |
| HC21 | - | - | - |
| HC22 | - | - | - |
| HC23 | - | - | - |
| HC24 | - | - | - |
| HC25 | - | - | - |
| HC26 | - | - | - |
| HC27 | - | - | - |
| HC28 | - | - | - |
| HC29 | - | - | - |
| HC30 | - | - | - |
| HC31 | - | - | - |
| HC32 | - | - | - |
| HC33 | - | - | - |
| HC34 | - | - | - |
| HC35 | - | - | - |
| HC36 | - | - | - |
| HC37 | - | - | - |
| HC38 | - | - | - |
| HC39 | - | - | - |
| HC40 | - | - | - |
| HC41 | - | - | - |
| HC42 | - | - | - |
| HC43 | - | - | - |
| HC44 | - | - | + |
| HC45 | - | - | - |
| HC46 | - | - | - |
| HC47 | - | - | - |
| HC48 | - | - | - |
| HC49 | - | - | - |
| HC50 | - | - | - |
| HC51 | - | - | - |
| HC52 | - | - | - |
| HC53 | - | + | - |
| HC54 | - | - | - |
| HC55 | - | + | - |
| HC56 | - | + | - |
| HC57 | - | - | - |
| HC58 | - | - | - |
| HC59 | - | - | - |
| HC60 | - | - | - |
| HC61 | - | - | - |

Antibody responses with SARS-CoV S1 and MERS-CoV S1 and S2 proteins were expressed in healthy controls. Data presented as positive (+) or negative (-).

**Extend Data Table 5.** Autoimmune antibodies in recovered COVID-19 individuals.

| Patient ID | ANA titer | Anti-ds-DNA ( IU/ml ) | Nucleosomes | Histones | SmD1 | U1-SnRNP | SS-A/Ro (60 kDa) | SS-A/Ro (52 kDa) | SS-B/La | Scl-70 | CENP-B | jo-1 | Anti-P0 (38 kDa) |
| --- | --- | --- | --- | --- | --- | --- | --- | --- | --- | --- | --- | --- | --- |
| PT1 | 1:20 | 28 | - | - | - | - | + | - | - | - | - | - | - |
| PT2 | <1:20 | 38 | - | - | + | - | - | - | - | - | - | - | - |
| PT3 | <1:20 | 27 | - | - | - | - | - | - | - | - | - | - | - |
| PT4 | <1:20 | 103 | - | - | - | - | - | - | - | - | - | - | - |
| PT5 | <1:20 | 35 | - | - | - | - | - | - | + | - | - | - | - |
| PT6 | <1:20 | 64 | - | - | - | - | + | - | - | - | - | - | - |
| PT7 | <1:20 | 17 | - | - | - | - | - | - | - | - | - | - | - |
| PT8 | <1:20 | 54 | - | - | - | - | - | - | - | - | - | - | - |
| PT9 | <1:20 | 43 | - | - | - | - | - | - | - | - | - | - | - |
| PT10 | <1:20 | 24 | - | - | - | - | - | - | - | - | - | - | - |
| PT11 | <1:20 | 55 | - | - | - | - | - | - | - | - | - | - | - |
| PT12 | 1:20 | 37 | - | - | - | - | - | - | - | - | - | - | - |
| PT13 | <1:20 | 88 | - | - | - | - | - | - | + | - | - | - | - |
| PT14 | <1:20 | 206 | - | - | - | - | - | - | - | - | - | - | - |
| PT15 | <1:20 | 31 | - | - | - | - | - | - | - | - | - | - | - |
| PT16 | <1:20 | 170 | - | - | - | - | - | - | - | - | - | - | - |
| PT17 | <1:20 | 23 | - | - | - | - | - | - | - | - | - | - | - |
| PT18 | <1:20 | 64 | - | - | - | - | - | - | + | - | - | - | - |
| PT19 | <1:20 | 35 | - | - | - | - | - | - | - | - | - | - | - |
| PT20 | <1:20 | 84 | - | - | - | - | - | - | - | - | - | - | - |
| PT21 | <1:20 | 50 | - | - | - | - | - | - | - | - | - | - | - |
| PT22 | <1:20 | 36 | - | - | - | - | - | - | - | - | - | - | - |
| PT23 | <1:20 | 46 | - | - | - | - | - | - | - | - | - | - | - |
| PT24 | <1:20 | 15 | - | - | - | - | - | - | - | - | - | - | - |
| PT25 | 1:40 | 28 | - | - | - | - | - | - | - | - | - | - | - |
| PT26 | <1:20 | 40 | - | - | - | - | - | - | - | - | - | - | - |
| PT27 | <1:20 | 43 | - | - | - | - | - | - | - | + | - | - | - |
| PT28 | <1:20 | 33 | - | - | - | - | - | - | - | - | - | - | - |
| PT29 | <1:20 | 39 | - | - | - | - | - | - | - | - | - | - | - |
| PT30 | <1:20 | 44 | - | - | - | - | - | - | - | - | - | - | - |
| PT31 | 1:20 | 63 | - | - | - | - | - | - | - | - | - | - | - |
| PT32 | <1:20 | 51 | - | - | - | - | - | - | - | - | - | - | - |
| PT33 | <1:20 | 30 | - | - | - | - | - | - | - | - | - | - | - |
| PT34 | 1:80 | 51 | - | - | - | - | + | - | - | - | - | - | - |
| PT35 | <1:20 | 25 | - | - | - | - | - | - | - | - | - | - | - |
| PT36 | <1:20 | 52 | - | - | - | - | - | - | - | - | - | - | - |
| PT37 | <1:20 | 46 | - | - | - | - | - | - | - | - | - | - | - |
| PT38 | <1:20 | 19 | - | - | - | - | - | - | - | - | - | - | - |
| PT39 | <1:20 | 35 | - | - | + | - | - | - | - | - | - | - | - |
| PT40 | <1:20 | 30 | - | - | - | - | - | - | - | - | - | - | - |
| PT41 | <1:20 | 17 | - | - | - | - | - | - | - | - | - | - | - |
| PT42 | <1:20 | 98 | - | - | - | - | - | - | - | - | - | - | - |
| PT43 | <1:20 | 15 | - | - | - | - | + | - | + | - | - | - | - |
| PT44 | <1:20 | 152 | - | - | - | - | - | - | - | - | - | - | - |
| PT45 | <1:20 | 28 | - | - | - | - | - | - | - | - | - | - | - |
| PT46 | <1:20 | 22 | - | - | - | - | - | - | - | - | - | - | - |
| PT47 | <1:20 | 24 | - | - | - | - | - | - | - | - | - | - | - |
| PT48 | <1:20 | 58 | - | - | - | - | - | - | - | - | - | - | - |
| PT49 | <1:20 | 37 | - | - | - | - | - | - | - | - | - | - | - |
| PT50 | <1:20 | 43 | - | - | - | - | - | - | - | - | - | - | - |
| PT51 | 1:80 | 50 | - | - | - | - | + | - | - | - | - | - | - |
| PT52 | 1:20 | 15 | - | - | - | - | - | - | - | - | - | - | - |
| PT53 | <1:20 | 37 | - | - | - | - | - | - | - | - | - | - | - |
| PT54 | <1:20 | 81 | - | - | - | + | - | - | - | - | - | - | - |
| PT55 | <1:20 | 34 | - | - | - | - | - | - | - | - | - | - | - |
| PT56 | <1:20 | 24 | - | - | - | - | - | - | - | - | - | - | - |
| PT57 | <1:20 | 43 | - | - | - | - | - | - | - | - | - | - | - |
| PT58 | <1:20 | 28 | - | - | - | - | - | - | - | - | - | - | - |
| PT59 | <1:20 | 30 | - | - | - | - | - | - | - | - | - | - | - |
| PT60 | <1:20 | 32 | - | - | - | + | - | - | - | - | - | - | - |
| **PT61** | <1:20 | 41 | - | - | - | - | - | - | - | - | - | - | - |
| **PT62** | <1:20 | 41 | - | - | - | - | - | - | - | - | - | - | - |
| **PT63** | <1:20 | 31 | - | - | - | - | - | - | - | - | - | - | - |
| **PT64** | <1:20 | 201 | - | - | - | - | - | - | - | - | - | - | - |
| **PT65** | <1:20 | 46 | - | - | - | - | - | - | - | - | - | - | - |
| **PT66** | <1:20 | 54 | - | - | - | - | - | - | - | - | - | - | - |
| **PT67** | <1:20 | 39 | - | - | - | - | - | - | - | - | - | - | - |
| ANA: Antinuclear antibodies; Anti-ds-DNA: Anti-double-stranded DNA; SmD1: Core small nuclear ribonucleoprotein particle splicing factor; SS-A/Ro: Anti-Sjogren's Syndrome antigen B antibody; SS-B/La: Anti-Sjogren's Syndrome B antibody; Scl-70: Autoantibodies to topoisomerase I; CENP-B: The major human centromere autoantigen; Jo-1: Histidyl-transfer RNA synthetase; Anti-P0: Anti-P antibodies react against acidic phosphorylated ribosomal proteins P0, P1, and P2 (with molecular mass of 38, 19, and 17 kDa, respectively). |  |  |  |  |  |  |  |  |  |  |  |  |  |

